# TMS-evoked phosphenes and oculomotor responses in visual-snow syndrome

**DOI:** 10.64898/2026.02.05.26344851

**Authors:** Roman Akinshin, Dinara Shankhoeva, Dmitry Gribanov, Artemiy Berkmush-Antipova, Polina Meschanina, Irina Tomilina, Elizaveta Ostrovskaia, Sergei Gostilovich, Nikolay Syrov, Anh-Huy Phan, Mikhail Lebedev

## Abstract

Visual Snow Syndrome (VSS) is a neurological condition characterized by continuous visual disturbances resembling television static across the visual field. Despite its significant impact on quality of life, objective assessment methods remain limited, with diagnosis relying primarily on subjective patient reports. Current understanding of VSS pathophysiology suggests cortical hyperexcitability, but precise mechanisms remain unclear. Here we developed an integrated protocol combining transcranial magnetic stimulation (TMS)-evoked phosphene mapping with high-resolution eye-tracking analysis. Four VSS patients diagnosed and eight age-matched healthy controls participated. Using individualized MRI-guided navigation, TMS pulses were delivered to occipital cortex regions. After each pulse, participants indicated perceived phosphene locations on a digital tablet. Eye movements were recorded continuously using a Tobii eye-tracker at 120 Hz. Participants completed prosaccade tasks to assess basic oculomotor function, and post-TMS oculomotor responses were analyzed. The VSS participants had significantly altered phosphene perception patterns with more diffuse spatial distribution compared to the controls’ more centralized and concentrated patterns. Prosaccade latencies were significantly shorter in VSS participants compared to healthy controls. Post-TMS oculomotor responses showed markedly reduced saccadic velocity in the VSS patients versus controls. Thus, the combination of phosphene mapping and eye-tracking analysis provides an efficient tool for VSS assessment and paves the way to uncovering its underlying pathophysiology.

## Introduction

VSS is a neurological perceptual disorder characterized by a persistent experience of fine, dynamic visual noise—typically described by patients as continuous “static” or flickering dots across the entire visual field [1]. The core phenomenon of visual snow is frequently accompanied by additional complaints, including palinopsia (afterimages or trailing), photophobia, nyctalopia, and enhanced entoptic phenomena (e.g., floaters) [1, 2, 3]. Non-visual symptoms such as tinnitus and a high prevalence of migraine are commonly reported comorbidities and contribute substantially to reduced quality of life in affected individuals [1, 2]. Routine ophthalmological examinations are generally unremarkable in VSS, suggesting a central (brain-based) origin of the syndrome [2].

Current mechanistic accounts of VSS converge on abnormalities in cortical excitability and large-scale brain networks. A widely discussed hypothesis posits occipital cortical hyperexcitability: electrophysiological studies report loss of normal habituation in visual evoked potentials and alterations that have been interpreted as increased visual-cortex responsiveness in some cohorts [2]. Neuroimaging investigations have also identified focal increases in metabolism and cerebral perfusion within occipital areas (including the lingual gyrus and adjacent visual cortices) in VSS patients, consistent with altered basal activity in early visual regions [4]. Complementary evidence from electroencephalography indicates disturbed rhythmic activity, such as alterations in occipital alpha and enhanced gamma-band responses – findings compatible with an imbalance of excitation and inhibition within visual circuits [3, 5]. Beyond focal cortical changes, structural and functional studies reveal broader network disturbances: altered connectivity between visual cortex and associative, attentional, and thalamic structures supports the view of VSS as a distributed network disorder with impaired sensory filtering and attentional gating [1, 4]. Thalamocortical dysrhythmia has been invoked as a possible unifying mechanism linking thalamic gating abnormalities with persistent cortical percepts in VSS, although direct evidence for this model remains under development [1]. Distinct, partially overlapping neural networks mediate perceptual and oculomotor functions relevant to VSS, with separate mechanisms governing sensory attention and motor planning within the parietal cortex [6]. These functional distinctions highlight the importance of investigating both perceptual experience and eye movement control jointly when studying VSS. Reviews also emphasize that despite advances in characterizing cortical hyperexcitability and network dysrhythmias, objective physiological biomarkers remain limited [7]. This underscores the need for multimodal paradigms capable of investigating network dysfunction in VSS.

Neuroimaging and electrophysiological tools (fMRI, PET, EEG/MEG, VEP) have advanced our understanding of VSS but have intrinsic limitations for linking regional neural dynamics to momentary perceptual experience. fMRI and PET provide useful spatial localization of altered activity and perfusion in VSS [4], yet their haemodynamic measures lack the temporal fidelity needed to resolve fast perceptual events. EEG and MEG capture millisecond-scale dynamics and reveal abnormal oscillatory patterns in VSS [3, 5], but their spatial precision and the correlational nature of these measures restrict causal inference. Visual evoked-potential paradigms highlight abnormal processing (e.g., poor habituation), yet they do not map perceptual experience retinotopically nor do they directly perturb cortical circuits to test causality. Despite these methodological limitations, convergent findings from multiple neuroimaging modalities have begun to establish a consistent neurobiological profile of VSS. Simultaneous 18F-FDG PET/MRI investigations, while limited in temporal resolution, demonstrate reproducible focal hypermetabolism in the right lingual gyrus and cuneus, with metabolic increases reaching 24% compared to healthy controls. This hypermetabolism co-localizes with structural grey matter volume increases at the lingual-fusiform junction, suggesting both functional hyperexcitability and anatomical alterations contribute to VSS pathophysiology [8]. While these converging multimodal findings establish a consistent neurobiological profile of VSS characterized by hyperexcitability and network dysfunction, the inherently correlational nature of neuroimaging and electrophysiological approaches limits their capacity to establish causal relationships between activity in specific visual cortical loci and the subjective visual symptoms of VSS [9].

TMS is a direct, noninvasive method to evaluate occipital cortex excitability and perception. Single TMS pulses applied over visual cortex elicit phosphenes in around ∼75% of the individuals—brief illusory flashes or shapes experienced in the absence of external visual input—which are caused by the activation of the retinotopically organized visual-cortex circuits [9, 10, 11]. The phosphene threshold (PT), defined as the minimum stimulation intensity required to evoke a percept on a defined proportion of trials, is widely used as a quantitative marker of local cortical excitability [12, 13]. Foundational work has mapped the retinotopic topography of TMS-evoked phosphenes and demonstrated systematic correspondences between coil position and perceived phosphene location in visual space [10, 11, 14]. Methodological refinements now permit precise mapping of TMS-evoked phosphenes, and perceived coordinates depend on the instantaneous gaze position, reflecting the retinotopic cortical organization and enabling reproducible mapping across trials and participants [15, 16]. Phosphene measures have shown test–retest reliability when rigorous protocols are applied, making them suitable for group comparisons and longitudinal study designs [14, 17].

Eye-tracking offers a measure of oculomotor control, which is tightly coupled to visual processing and attention. High-resolution video-based trackers quantify saccades, microsaccades, fixation stability, and pursuit with millisecond timing and are widely used to reveal subtle sensory–motor dysfunction. In VSS, eye-tracking studies report consistent alterations of oculomotor behaviour: shortened prosaccade latencies, increased antisaccade error rates, impaired fixation stability, and abnormalities in reading-related eye movements have all been described, suggesting changes in the balance of reflexive and voluntary oculomotor control and attentional selection processes [18, 19, 20]. Because eye movements both reflect and influence perceptual processing (for example, saccades transiently modulate visual sensitivity), combining gaze metrics with direct cortical perturbation can reveal mechanistic links between perception and motor output [21].

Integrating TMS-evoked phosphene mapping with synchronous eye-tracking therefore offers a powerful, causal framework to study VSS. In a combined paradigm, MRI-guided TMS targets retinotopically defined occipital loci while eye position is monitored continuously: subjects report phosphene attributes and locations (e.g., by marking perceived positions on a calibrated grid) and eye-tracking captures any reflexive gaze shifts or micro-saccadic adjustments time-locked to stimulation. This approach simultaneously yields spatially resolved maps of cortical excitability (phosphene probability, location, and threshold) and temporally precise measures of oculomotor response to cortical activation. These dimensions complement one another: phosphene topography reveals where cortical processing is altered, while gaze dynamics indicate how oculomotor circuitry reacts to that alteration. Although TMS-phosphene methods and perceptual mapping have been applied in healthy vision research [14, 15, 16] and oculomotor profiling has been used in VSS cohorts [17, 18], a systematic application of combined TMS and high-resolution eye-tracking specifically in VSS cohorts has not been conducted.

The design of this study included participant selection, MRI-guided TMS targeting, phosphene-mapping procedures, synchronized eye-tracking acquisition, and analysis pipelines developed to quantify phosphene topography and post-stimulation oculomotor dynamics. With this methodology, we mapped perceptual sensitivity across visual field locations in VSS, quantified altered cortical excitability via phosphene thresholds and distributions, and characterized the oculomotor signatures associated with thalamocortical dysrythmias.

## Methods

### Participants

Twelve right-handed individuals took part in the study: four patients with VSS (mean age 27.3 ± 2.6 years, 2 females) and eight healthy controls (HC; mean age 25.1 ± 3.2 years, 4 females). VSS was diagnosed by a neurologist according to the ICHD-3 criteria, and none of the controls reported symptoms of persistent visual snow, migraine, or other neurological conditions. Exclusion criteria included contraindications to transcranial magnetic stimulation (TMS; e.g., epilepsy, metallic implants, pacemakers), history of psychiatric disorders, or uncorrected visual impairment. All participants had normal or corrected-to-normal vision. Written informed consent was obtained from each participant, and the study was approved by the Ethics Committee of Skoltech University, in accordance with the Declaration of Helsinki. The study was registered as a clinical trial at ClinicalTrials.gov (NCT07110493).

### Equipment

Stimulation was delivered using a Nexstim Navigated Brain Stimulation (NBS) system equipped with a figure-of-eight coil (70 mm). Frameless stereotactic MRI-guided navigation ensured accurate positioning of the coil over occipital cortex (Brodmann areas 17–19). Individual high-resolution anatomical MRIs were used when available; otherwise, a standardized MNI-space template was applied. Coil position and orientation were continuously tracked and logged by the NBS system with submillimeter accuracy.

Eye movements were recorded using a Tobii Pro video-based eye-tracker sampling at 120 Hz. Calibration was performed at the beginning of each session with a standard nine-point procedure. Eye-tracking data streams were time-locked to TMS pulses using shared digital triggers.

Phosphene locations were marked by participants on a Wacom Intuos graphics tablet with a superimposed coordinate grid covering ±20° of visual angle. The tablet output was synchronized to the shared experimental clock and stored with time-stamps for each response

## Procedure

### Phosphene threshold determination

At the start of Session 1, the individual phosphene threshold (PT) was determined. PT was defined as the lowest percentage of maximum stimulator output (MSO) that elicited a phosphene in at least 5 of 10 consecutive single-pulse trials delivered over occipital cortex during central fixation.

### Phosphene mapping protocol

Once PT was established, single TMS pulses at PT intensity were delivered at multiple occipital scalp sites. Each stimulation event was logged with a timestamp, which was synced with the eye-tracking data and tablet input. This ensured precise temporal alignment across all modalities. After each stimulation, participants localized the perceived phosphene by drawing its position and approximate shape on the digital grid. Each site was stimulated 10 times. Eye-tracking data were analyzed in the 0–500 ms post-stimulus window, defined relative to the timestamp of the TMS pulse, to capture oculomotor responses.

**Fig. 1.**
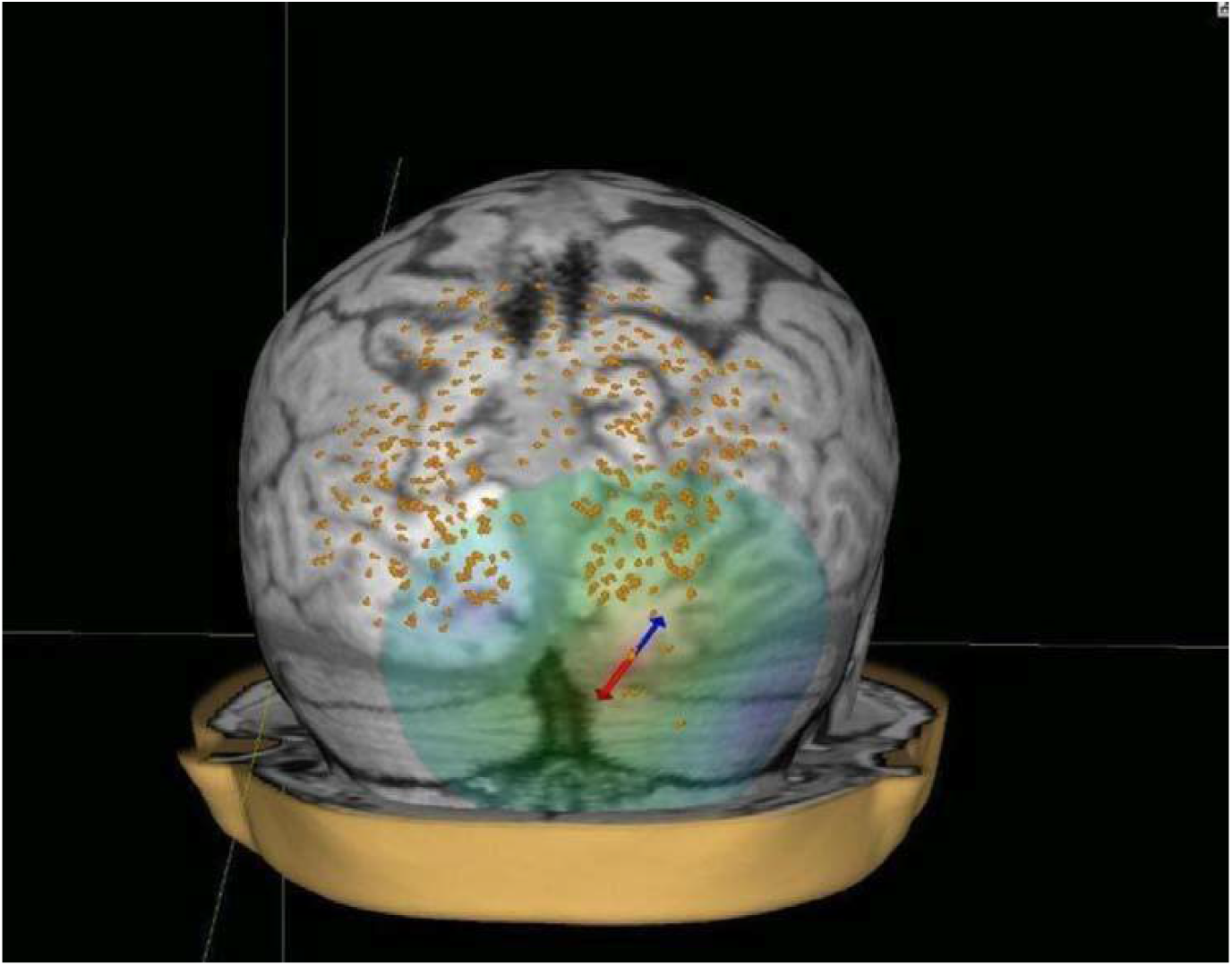
A map of cortical locations where TMS evoked phosphenes.

### Oculomotor task session

In a separate session, participants completed a psychomotor task (prosaccade paradigm). A central fixation cross was presented, followed by a peripheral target (10° left or right). Participants were instructed to look at the target as quickly and accurately as possible. Stimulus onset was logged with millisecond precision, and eye traces were synced with the presentation clock.

## Data Analysis

All data streams (TMS triggers, coil coordinates, eye-tracking traces, and tablet inputs) were merged using shared time-stamps. This ensured temporal alignment of stimulation events, perceptual reports, and oculomotor responses.

### Phosphene data collection

Tablet responses were digitized and processed using Python and OpenCV. For each drawing, the center of mass was extracted and mapped into normalized visual field coordinates. Individual subject maps were constructed by aggregating trial-wise phosphene centers into two-dimensional occurrence histograms. Group-level maps were obtained by averaging across participants in each cohort (HC and VSS).

Mean PT (expressed as % of maximum stimulator output) was calculated per subject. Group-level comparisons (VSS vs HC) were performed using independent-samples t-tests.

### Oculomotor responses immediately following TMS

For each phosphene-evoking trial, eye position was segmented into a 0–500 ms epoch following the TMS pulse timestamp. Gaze velocity (px/s) was computed as the derivative of horizontal and vertical position traces. The primary dependent variable was mean gaze velocity across the epoch. Participant-level averages were compared between groups using t-tests.

### Prosaccades

For the prosaccade task, latency (time from target onset to gaze initiation, relative to stimulus timestamps) and peak velocity (maximum instantaneous gaze speed reached during the execution of a saccade) were computed. Latency distributions were analyzed per subject and then aggregated by group. Between-group differences were assessed with t-tests.

## Results

### Phosphene thresholds

VSS participants exhibited significantly lower phosphene thresholds than controls (46.2 ± 5.7% MSO vs 59.1 ± 6.3% MSO; t = 3.97, p < 0.01). This indicated increased excitability of the occipital cortex in VSS.

### Spatial distributions of phosphenes

Control participants produced compact, retinotopically consistent phosphene fields, typically located near the foveal region. In contrast, VSS participants reported broader, more diffuse phosphenes with greater trial-to-trial variability.

**Figure 2.**
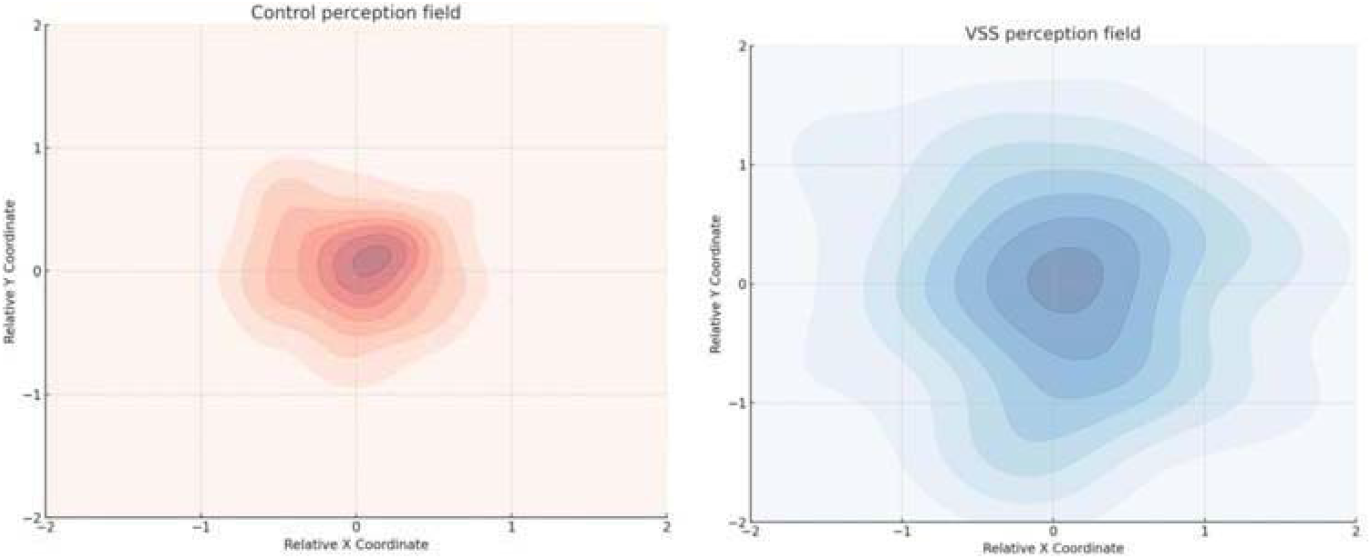
Group-level heatmaps of phosphene distributions in HC (left) and VSS (right).

### Oculomotor responses immediately following TMS

During the 500-ms interval following a TMS pulse, mean gaze velocity was markedly reduced in VSS (0.61 ± 0.13 px/s) compared to HC (2.84 ± 0.21 px/s; t = 14.49, p < 0.001). This suggests an impaired translation of cortical activation into an oculomotor response.

**Figure 3.**
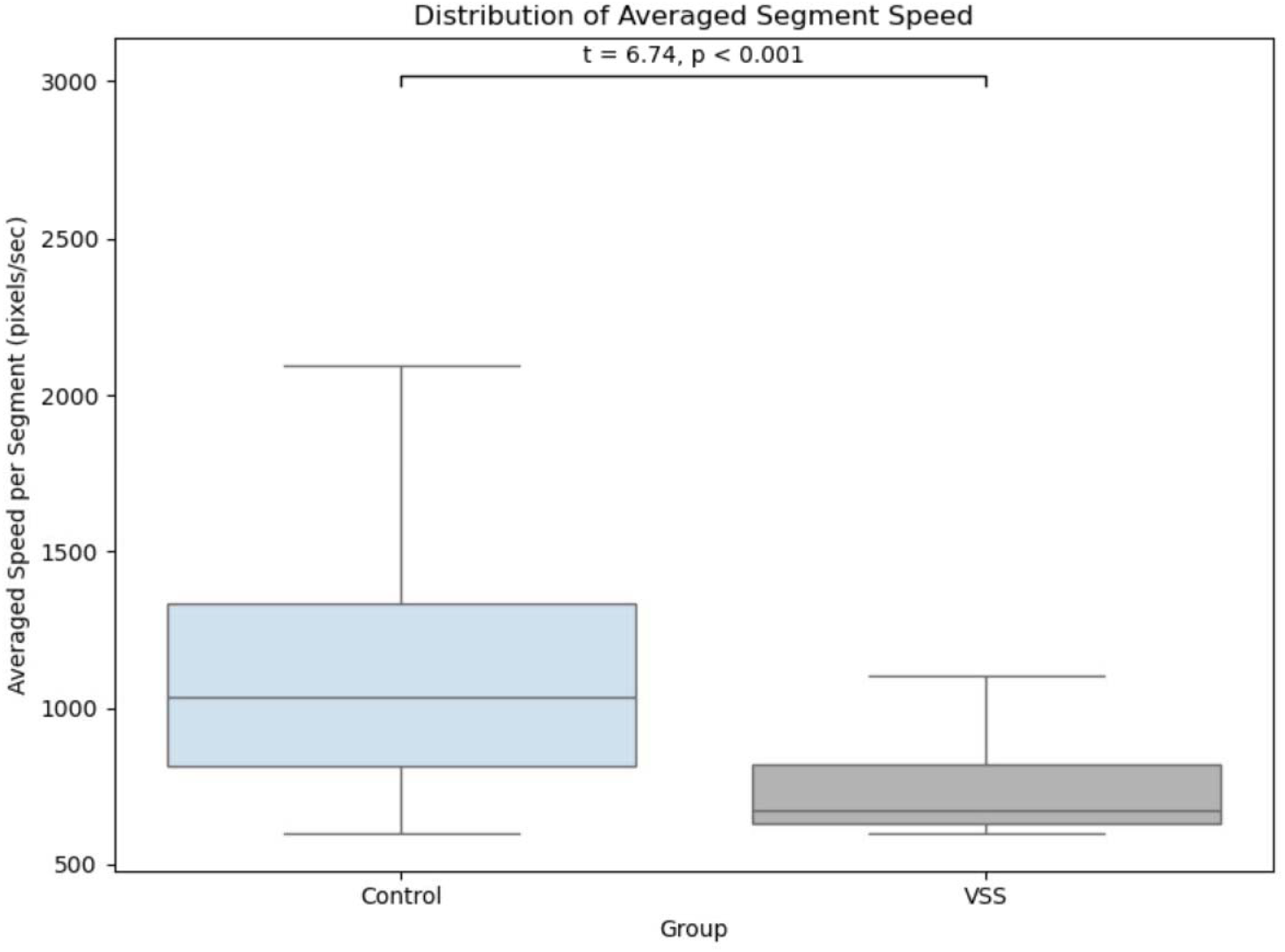
Distribution of average speed (pixels/s) for the 500-ms interval immediately following a TMS pulse for the HC and VSS groups. The central horizontal line indicates the median, the box represents the interquartile range (IQR), and whiskers extend to 1.5 times the IQR. The bar above the boxplots indicates a significant difference between groups (t-test: t = 6.74, p < 0.001)

### Prosaccades

VSS participants showed shorter saccade latencies (215.9 ± 22.4 ms) than HC (251.4 ± 28.6 ms; t = 4.29, p = 0.0046), while peak saccade velocity did not differ significantly between the groups.

**Figure 4.**
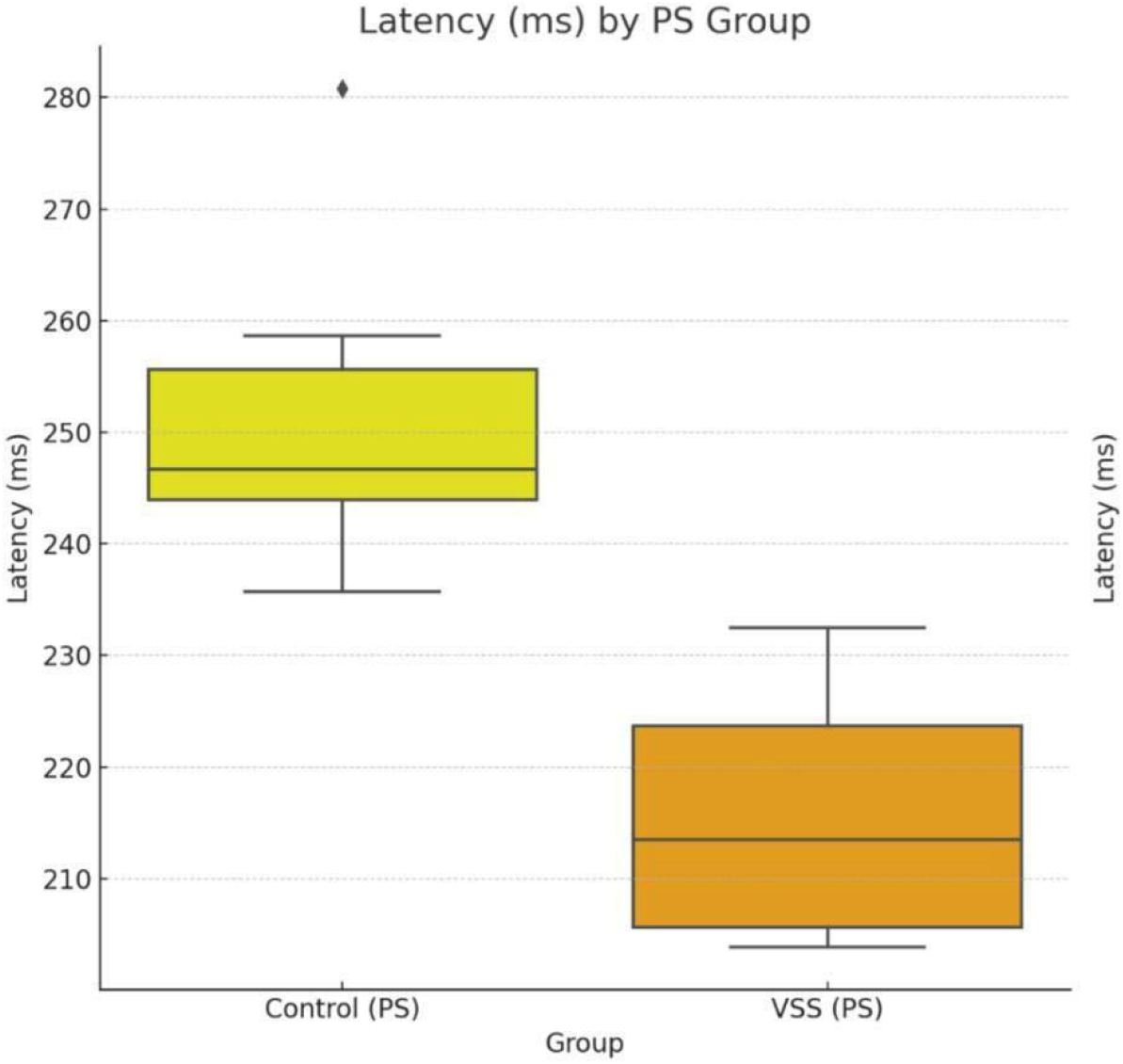
Boxplot of prosaccade latencies in VSS vs HC

## Discussion

In this study, we employed a methodology for the assessment of TMS-evoked phosphenes and oculomotor dynamics in individuals with Visual Snow Syndrome (VSS) compared to healthy controls (HC). By combining MRI-guided TMS with synchronized eye-tracking and digital phosphene mapping, we directly quantified both cortical excitability and visuomotor responses using a unified experimental framework. The present findings provide convergent evidence that VSS is characterized by heightened cortical excitability, diffuse perceptual mapping, and altered translation of TMS-evoked cortical activation into oculomotor output.

### Phosphene thresholds and cortical excitability

We observed significantly reduced phosphene thresholds in VSS relative to controls, indicating increased excitability of visual cortex. This is consistent with prior reports of reduced phosphene thresholds and impaired habituation of visual evoked potentials in VSS [2], as well as neuroimaging evidence of increased perfusion and metabolism in the lingual gyrus and other occipital regions [4]. The present data therefore reinforce the view that VSS involves a hyperexcitable occipital cortex [2, 22]. Importantly, phosphene thresholds provide a direct, causal readout of local excitability, in contrast to the correlational nature of neuroimaging and electrophysiology.

### Diffuse phosphene spatial distributions

Phosphene maps in HC were compact and retinotopically stable, whereas VSS participants produced broader, more variable distributions. This diffuse pattern could reflect a larger population of occipital neurons being recruited by a single TMS pulse, potentially due to impaired inhibitory mechanisms in early visual cortex [1]. This interpretation is supported by electrophysiological findings of abnormal oscillatory activity in VSS, including reduced alpha modulation [5] and enhanced gamma-band synchronization [3]. A loss of spatial precision in cortical activation could contribute to the phenomenology of persistent “visual snow,” where irrelevant or noisy signals fail to be filtered from conscious perception [1].

### Oculomotor responses to TMS

Eye-tracking during the 500-ms interval following phosphene-inducing TMS pulses revealed between-group differences. Healthy controls exhibited rapid gaze adjustments with higher mean velocities, whereas VSS patients had significantly reduced post-stimulus gaze speed. This impairment occurred within a reflexive time window, indicating disrupted translation of cortical activation into oculomotor output rather than deliberate compensatory behavior. These findings align with earlier work showing abnormalities in fixation stability and pursuit in VSS [18, 19], and extend them by demonstrating causal disruption of visuomotor coupling following direct cortical stimulation.

Taken together, reduced thresholds, diffuse phosphene fields, and weakened post-TMS oculomotor responses converge on a model of VSS as a disorder of cortical excitability and network-level dysregulation [1, 4, 14]. Thalamocortical dysrhythmia [18] and impaired sensory filtering [1] have been proposed as candidate mechanisms, and our findings provide experimental support for both: heightened occipital excitability on the one hand, and reduced fidelity of perceptual–motor coupling on the other. Importantly, this dual-modality paradigm linking cortical activation (phosphenes) to immediate motor consequences (gaze dynamics) offers a novel framework for probing causal mechanisms in VSS, beyond correlational imaging approaches.

### Methodological contribution

This study demonstrates the efficiency of combining TMS-induced phosphenes with synchronized eye-tracking as an objective tool for assessing visuoperceptual and oculomotor dynamics in VSS. The approach integrates spatial mapping, excitability thresholds, and behavioral coupling into a single experimental protocol. While further validation in larger samples will be needed, the current data highlight the potential of this paradigm as a biomarker framework for disorders of visual processing, including VSS and related conditions such as migraine [22, 23].

## Conclusion

This study introduces and validates a novel methodology for the assessment of TMS-evoked phosphenes and oculomotor dynamics in VSS. By integrating MRI-guided TMS with synchronized eye-tracking and digital phosphene mapping, we demonstrate that VSS is characterized by reduced phosphene thresholds, diffuse and unstable perceptual fields, and altered visuomotor responses to cortical stimulation. These findings provide causal evidence of abnormal cortical excitability and visuomotor integration in VSS. Beyond characterizing disease-specific alterations, the proposed paradigm establishes a methodological framework for linking cortical activation to perceptual and motor outcomes in real time. This approach may serve as a foundation for the development of objective biomarkers and translational tools for VSS and related disorders of visual perception.

## Data Availability

All data produced in the present study are available upon reasonable request to the authors

## References

1. Klein, A., & Schankin, C. J. (2021). Visual Snow Syndrome as a network disorder: A systematic review. Frontiers in Neurology, 12. 10.3389/fneur.2021.724072

2. Yildiz, F. G., Turkyilmaz, U., & Unal Cevik, I. (2019). The clinical characteristics and neurophysiological assessments of the occipital cortex in visual snow syndrome with or without migraine. Headache: The Journal of Head and Face Pain, 59(4), 484–494. 10.1111/head.13494

3. Orekhova, E. V., Naumova, S. M., Obukhova, T. S., Plieva, A. M., Prokofiev, A. O., Petrokovskaia, A. V., Artemenko, A. R., & Stroganova, T. A. (2025). Enhanced Neural Plasticity of the Primary Visual Cortex in Visual Snow Syndrome: Evidence from Meg Gamma Oscillations. 10.1101/2025.02.12.637794

4. Puledda, F., Schankin, C. J., O’Daly, O., Ffytche, D., Eren, O., Karsan, N., Williams, S. C., Zelaya, F., & Goadsby, P. J. (2021). Localised increase in regional cerebral perfusion in patients with visual snow syndrome: A pseudo-continuous arterial spin labelling study. Journal of Neurology, Neurosurgery & Psychiatry, 92(9), 918–926. 10.1136/jnnp-2020-325881

5. Klein, A., Aeschlimann, S. A., Zubler, F., Scutelnic, A., Riederer, F., Ertl, M., & Schankin, C. J. (2024). Alterations of the alpha rhythm in visual snow syndrome: A case-control study. The Journal of Headache and Pain, 25(1). 10.1186/s10194-024-01754-x

6. Huddleston, W. E., Swanson, A. N., Lytle, J. R., & Aleksandrowicz, M. S. (2021). Distinct saccade planning and endogenous visuospatial attention maps in parietal cortex: A basis for functional differences in sensory and motor attention. Cortex, 137, 292–304. 10.1016/j.cortex.2021.01.009

7. Rusztyn, P., Stańska, W., Torbus, A., & Maciejewicz, P. (2023). Visual snow: A review on pathophysiology and treatment. Journal of Clinical Medicine, 12(12), 3868. 10.3390/jcm12123868

8. Van Laere, K., Ceccarini, J., Gebruers, J., Goffin, K., & Boon, E. (2022). Simultaneous 18F-FDG PET/MR metabolic and structural changes in visual snow syndrome and diagnostic use. EJNMMI research, 12(1), 77. 10.1186/s13550-022-00949-0

9. Hallett, M. (2000). Transcranial magnetic stimulation and the human brain. Nature, 406(6792), 147–150. 10.1038/35018000

10. Kammer, T. (1998). Phosphenes and transient scotomas induced by magnetic stimulation of the occipital lobe: Their topographic relationship. Neuropsychologia, 37(2), 191–198. 10.1016/s0028-3932(98)00093-1

11. Tani, N., Hirata, M., Motoki, Y., Saitoh, Y., Yanagisawa, T., Goto, T., Hosomi, K., Kozu, A., Kishima, H., & Yorifuji, S. (2011). Quantitative analysis of phosphenes induced by navigation-guided repetitive transcranial magnetic stimulation. Brain Stimulation, 4(1), 28–37. 10.1016/j.brs.2010.03.006s

12. Boroojerdi, Babak, Meister, I. G., Foltys, H., Sparing, R., Cohen, L. G., & Töpper, R. (2002). Visual and motor cortex excitability: A transcranial magnetic stimulation study. Clinical Neurophysiology, 113(9), 1501–1504. 10.1016/s1388-2457(02)00198-0

13. Boroojerdi, B., Prager, A., Muellbacher, W., & Cohen, L. G. (2000). Reduction of human visual cortex excitability using 1-Hz transcranial magnetic stimulation. Neurology, 54(7), 1529–1531. 10.1212/wnl.54.7.1529

14. Elkin-Frankston, S., Fried, P. J., Pascual-Leone, A., Rushmore III, R. J., & Valero-Cabré, A. (2010). A novel approach for documenting phosphenes induced by transcranial magnetic stimulation. Journal of Visualized Experiments, (38). 10.3791/1762

15. Silva, A. E., Tsang, K., Hasan, S. J., & Thompson, B. (2021). Precise oculocentric mapping of transcranial magnetic stimulation-evoked phosphenes. NeuroReport, 32(11), 913–917. 10.1097/wnr.0000000000001683

16. Schaeffner, L. F., & Welchman, A. E. (2016). Mapping the visual brain areas susceptible to phosphene induction through brain stimulation. Experimental Brain Research, 235(1), 205–217. 10.1007/s00221-016-4784-4

17. Siniatchkin, M., Schlicke, C., & Stephani, U. (2011). Transcranial magnetic stimulation reveals high test–retest reliability for phosphenes but not for suppression of visual perception. Clinical Neurophysiology, 122(12), 2475–2481. 10.1016/j.clinph.2011.05.003

18. Solly, E. J., Clough, M., McKendrick, A. M., Foletta, P., White, O. B., & Fielding, J. (2021). Eye Movement characteristics provide an objective measure of visual processing changes in patients with visual snow syndrome. Scientific Reports, 11(1). 10.1038/s41598-021-88788-2

19. Solly, E. J., Clough, M., McKendrick, A. M., Foletta, P., White, O. B., & Fielding, J. (2020). Ocular motor measures of visual processing changes in visual snow syndrome. Neurology, 95(13). 10.1212/wnl.0000000000010372

20. Tannen, B., Sample, A., Ciuffreda, K. J., & Tannen, N. M. (2024). Clinical reading-related oculomotor assessment in visual snow syndrome. Journal of Optometry, 17(2), 100500. 10.1016/j.optom.2023.100500

21. Goettker, A., Braun, D. I., Schütz, A. C., & Gegenfurtner, K. R. (2018). Execution of saccadic eye movements affects speed perception. Proceedings of the National Academy of Sciences, 115(9), 2240–2245. 10.1073/pnas.1704799115

22. Ekkert, A., Noreikaitė, K., Valiulis, V., & Ryliškienė, K. (2019). Migraine-linked characteristics of transcranial magnetic stimulation-induced phosphenes. Journal of Integrative Neuroscience, 18(4). 10.31083/j.jin.2019.04.1182

23. Kammer, T., Puls, K., Erb, M., & Grodd, W. (2004). Transcranial magnetic stimulation in the visual system. II. characterization of induced phosphenes and scotomas. Experimental Brain Research, 160(1), 129–140. 10.1007/s00221-004-1992-0

